# Impact of increasing CD4 count threshold eligibility for antiretroviral therapy initiation on advanced HIV disease and tuberculosis prevalence and incidence in South Africa: an interrupted time series analysis

**DOI:** 10.1101/2024.06.21.24309333

**Authors:** Kwabena Asare, Lara Lewis, Johan van der Molen, Yukteshwar Sookrajh, Thokozani Khubone, Thulani Ngwenya, Mkhize Siyabonga Ntokozo, Richard J Lessells, Kogieleum Naidoo, Phelelani Sosibo, Christian Bottomley, Nigel Garrett, Jienchi Dorward

## Abstract

**Introduction:** We investigated the impact of increasing CD4 count eligibility for antiretroviral-therapy (ART) initiation on advanced HIV and tuberculosis (TB) prevalence and incidence among people living with HIV (PLHIV) in South Africa.

**Methods:** We conducted an interrupted time series analysis with de-identified data of PLHIV aged ≥15 initiating ART between April-2012 and February-2020 at 65 primary healthcare clinics in KwaZulu-Natal, South Africa. Outcomes included monthly proportions of new ART initiators presenting with advanced HIV (CD4 count <200 cells/µl) and TB disease. We created a cohort of monthly ART initiators without TB and evaluated the cumulative incidence of TB within 12 months follow-up. We used segmented binomial regression models to estimate relative risks (RR) of outcomes, allowing for a step and slope change after expanding the ART initiation CD4 count eligibility from <350 to <500 cells/µl in January- 2015 and following Universal-Test-and-Treat (UTT) implementation in September-2016.

**Results:** Among 187,544 participants, median age was 32 (27-39), and 125,065 (66.7%) were female. After January-2015, risk of advanced HIV at initiation decreased by 24.5% (RR=0.745, 95%CI 0.690-0.800) and further reduced by 26.2% following UTT implementation (RR=0.738, 95%CI 0.688-0.788). Risk of TB at initiation also decreased by 28.7% after January-2015 (RR=0.713, 95%CI 0.644-0.782) and further decreased by 17.6% after UTT implementation (RR=0.824, 95%CI 0.703-0.945) but remained stable among initiators with advanced HIV. Among the incidence cohort, the risk of new TB decreased by 31.9% (RR=0.681, 95%CI 0.441-0.921) following UTT implementation. Among the incidence cohort with advanced HIV, there was weak evidence of a decrease in risk of new TB (RR=0.755, 95%CI 0.489-1.021), but it gradually decreased per month (slope change per month 9.7%, RR=0.903, 95%CI 0.872-0.934) following UTT implementation.

**Conclusions:** Our data supports the added benefit of decreased TB co-burden with expanded ART access. Early diagnosis and immediate linkage to care should be prioritised among PLHIV.

## INTRODUCTION

During the HIV epidemic, there has been a gradual expansion of the CD4 count eligibility criteria for initiating people living with HIV (PLHIV) on antiretroviral therapy (ART) in South Africa. Before August 2011, the initiation CD4 count eligibility was <200 cells/µl.[1] From August 2011, it was expanded to <350 cells/µl and then to <500 cells/µl from January 2015.[1] In September 2016, the World Health Organisation’s (WHO) Universal-Test-and- Treat (UTT) policy was implemented in South Africa, allowing ART initiations regardless of CD4 count level or clinical stage of HIV disease.[2, 3] Likewise, from April 2012, even before UTT, all PLHIV with active tuberculosis disease (TB) in South Africa became eligible for ART initiation, irrespective of the CD4 count level.[4] These policy changes were informed by evidence of the efficacy of early ART initiation on viral suppression[5] and the efficacy of combining ART with TB treatment in reducing all-cause mortality.[6]

After implementing these policies, access to ART increased, and a higher proportion of PLHIV in South Africa and other LMICs initiated ART at higher CD4 counts.[7–9] Consequently, a higher proportion of people receiving ART have achieved viral suppression[10, 11], and population-level HIV incidence has reduced.[12] The life expectancy of PLHIV has also improved[13], and the burden of opportunistic co-infections such as TB has reduced.[14] However, these positive health outcomes are likely due to more PLHIV starting ART earlier at higher CD4 counts due to the expansion of initiation CD4 count eligibility over time. A number of cohort and cross-sectional studies have demonstrated the positive health related impacts of expanded ART access[9, 14–18] in LMICs but there is limited evidence on the stepwise impact of ART eligibility expansion on trends in advanced HIV disease and the risk of common causes of death, such as TB disease in PLHIV initiating and receiving ART.

We aimed to determine the impact of increasing the ART initiation CD4 count eligibility from <350 to <500 cells/μl in January 2015 and then to UTT implementation in September 2016 on trends in advanced HIV disease at ART initiation, TB prevalence at ART initiation and cumulative TB incidence within 12 months follow-up. We aimed to evaluate these objectives among ART initiators and in a sub-cohort of ART initiators with advanced HIV disease from April 2012 to February 2020 in South Africa.

## METHODS

### Study design and setting

We conducted a segmented interrupted time series analysis with de-identified routinely collected data from 65 public primary healthcare facilities in KwaZulu-Natal, South Africa, from 2012 to 2020. Fifty-six clinics were in the eThekwini District, and the remaining 9 fixed and mobile primary care clinics in the rural uMkhanyakude district. Based on prevailing clinical guidelines, ART initiation was predicated by clinical assessment for pregnancy in females, and CD4 count testing and screening for TB disease.[19, 20] The guidelines recommend screening for TB and pregnancy in females during the follow-up clinic visits. TB screening involves assessments for TB symptoms followed with GeneXpert testing among symptomatic participants.[19–21] But all pregnant women are recommended to received GeneXpert testing regardless of TB symptoms, due to the lower sensitivity of the TB symptom screen in pregnant women.[19] TB preventive therapy (TPT) is recommended for participants for whom active TB is reasonably ruled out based on TB symptom screening.[19, 20] Follow-up clinic visits were usually between 1 to 3 months apart and viral load testing were usually performed 1 to 2 times yearly.

We selected the study period to start in April 2012, when PLHIV who had TB became eligible for ART initiation, regardless of CD4 count in South Africa.[4] We chose February 2020 as the study endpoint to exclude the COVID-19 lockdown period in South Africa, which started in March 2020.[22]

### Data sources and data management

We used data from South Africa’s TIER.Net electronic database containing demographic and clinical data on PLHIV receiving ART and TB care in public sector healthcare clinics.[23] The information captured includes ART and TB treatment dates and regimens, laboratory results including CD4 count, clinic visits and TB status at ART initiation and at each follow- up clinic visit.

### Participants

For the main cohort, we included all PLHIV aged ≥15 years initiating ART at the study clinics from April 2012 to February 2020. Additionally, we analyzed an incidence cohort comprising a sub-group of the main cohort who initiated ART without TB from April 2012 to February 2019. For the incidence cohort, the time frame of ART initiation was to allow a minimum of 365 days follow-up duration for all participants before the end of the study period in February 2020. Participants entered the incidence cohort at ART initiation and exited at the earliest of (1) outcome onset (i.e., status change to “on TB treatment” within 12 months), (2) end of the 12 months follow-up or (3) censored within 12 months (lost to follow-up, died, or transferred out as we could not access or link participants to data at other clinics). We also analyzed sub-cohorts of the main and incidence cohorts for each category of age (15-24, 25-34, 35-44, 45-54 and 55+ years), gender (Male and Female), and advanced HIV disease at initiation.

### Outcomes

The study outcomes were the monthly number of new ART initiators (main cohort), the monthly proportion of new ART initiators with advanced HIV disease (main cohort), the monthly number and proportion of new ART initiators with active TB disease (main cohort) and the number and proportion of monthly new ART initiators developing new active TB disease within 12 months follow-up (incidence cohort).

We estimated the monthly number of new ART initiators (excluding participants who were transferring in from another clinic) by counting the number of new ART initiation dates per month and, among these, the number with TB and the number with advanced HIV disease (i.e., having a CD4 count <200 cells/µl). TB at ART initiation meant the participant was on TB treatment at the start of ART. We calculated the monthly proportion of new ART initiators with advanced HIV disease as the number of new ART initiators with advanced HIV disease divided by the monthly number of new ART initiators. We calculated the monthly proportion of new ART initiators with TB as the number of new ART initiators with TB disease divided by the monthly number of new ART initiators. Thus, the monthly proportion of new ART initiators with TB includes participants with unascertained TB status and hence captures the proportion of initiators with a known TB status.

Among the incidence cohort, new TB meant that the participant’s status changed to “on TB treatment” within 12 months follow-up after ART initiation. In the incidence cohort, we counted the monthly number of ART initiators and of these counted the number that developed new TB within 12 months of follow-up (new TB cases). We estimated the proportion of new TB within 12 months as the number of new TB cases divided by the monthly number of ART initiators. Thus, the proportion of monthly ART initiators developing new TB includes participants with unascertained new TB status and hence captures the proportion of initiators with a known new TB status.

### Statistical analyses

We calculated descriptive summaries of participant demographics, clinic data and crude summaries of the primary outcomes from the main and incidence cohorts among participants initiating ART in each CD4 count eligibility period. The demographic variables included in the descriptive summaries were age, gender, pregnancy status in females and district settlement type of the clinics (rural uMkhanyakude and urban eThekwini). We included the most recent available CD4 count recorded from before 180 and up to 30 days after ART initiation.

We fitted segmented time series regression models of all outcomes. The time series started from a baseline period of April 2012 to December 2014 when the initiation CD4 count eligibility was <350 cells/μl and allowing for step and slope changes from January 2015 to capture the impact of expanding the initiation CD4 count eligibility to <500 cells/μl and from September 2016 to capture the impact of UTT implementation. We conducted linear regression of the monthly number of new ART initiators, monthly number of new ART initiators with TB and number of monthly new ART initiators developing new TB within 12 months follow-up using the Prais-Winsten method to account for serial autocorrelation of the errors.[24] We performed binomial regression of the monthly proportion of new ART initiators with advanced HIV disease, monthly proportion of new ART initiators with TB and proportion of monthly new ART initiators developing new TB within 12 months follow-up. We used a log link for the binomial regression models and exponentiated the coefficients to estimate relative risks (RR) and used Newey-West standard errors with lags up to 3 to calculate confidence intervals.[25, 26]

We included the following predictor variables in each model: a time variable measured in months (i.e., the month of ART initiation), dummy variables indicating pre or post implementation date of each CD4 count eligibility expansion (i.e., January 2015 for expansion to <500 cells/μl and September 2016 for UTT implementation) to capture the step change in outcomes after each date, a variable indicating time in months since January 2015 (zero for the months before) and time in months since September 2016 (zero for the months before) to capture the slope changes in outcomes, and 2 Fourier terms consisting of two pairs of sine and cosine transformations of a 12-month cycle to adjust for seasonal trends in outcomes.[27]

We plotted the segmented interrupted time series graphs from each regression model. We included the counterfactual scenarios for each model, defined as situations where only the previous initiation CD4 count eligibility criteria were implemented. The counterfactuals were estimated by predicting the outcomes from the segmented regression models, using a modified dataset as: (A) the pre-post January 2015 and pre-post September 2016 dummy variables = No and time since January 2015 and time since September 2016 variables = 0 to predict the counterfactuals from January 2015 to August 2016 and, (B) the pre-post September 2016 dummy variable = No and time since September 2016 variable = 0 to predict the counterfactuals after UTT implementation).

We conducted sub-cohort analyses of the segmented regression and interrupted time series by age category (15-24, 25-34, 35-44, 45-54 and 55+ years), gender (Male and Female), and among initiators with advanced HIV disease. We performed all statistical analyses using R 4.2.2 (R Foundation for Statistical Computing, Vienna, Austria).[28]

### Patients and public involvement

We used de-identified routinely collected data with no contacts to the patients. Hence patients and public involvement in the design, conduct, reporting or dissemination of our research was not applicable.

## RESULTS

### Characteristics of participants at ART initiation

From April 2012 to February 2020, 187,544 PLHIV aged ≥15 years initiated ART at the clinics (Table 1a). The median age was 32 years (interquartile range [IQR] 27-39). In total, 125,065 (66.7%) were female, of whom 24,447 (19.5%) were pregnant at the time of ART initiation. The median CD4 count at ART initiation per period was 227 cells/μl (IQR 130- 313) from April 2012 to December 2014, when the initiation CD4 count eligibility was <350 cells/μl, 306 cells/μl (IQR 176-425) from January 2015 to August 2016 when the initiation CD4 count eligibility was <500 cells/μl and 368 cells/μl (IQR 211-557) from September 2016 to February 2020 during UTT implementation. The number and proportion of initiators with CD4 count <200 cells/μl was 21,839 (42.6%) when the initiation CD4 count eligibility was <350 cells/μl, 13,700 (29.2%) when the initiation CD4 count eligibility was <500 cells/μl and 20,830 (23.3%) during UTT implementation. The number and proportion of initiators with TB were 7,835 (15.3%) when the initiation CD4 count eligibility was <350 cells/μl, 5,318 (11.3%) when the initiation CD4 count eligibility was <500 cells/μl and 6,911 (7.7%) during UTT implementation. Among initiators with advanced HIV disease, the number and proportion with TB was 4,538 (20.8%) when the initiation CD4 count eligibility was <350 cells/μl, 2,875 (21.0%) when the initiation CD4 count eligibility was <500 cells/μl and 3,839 (18.4%) during UTT implementation.

**Table 1.**
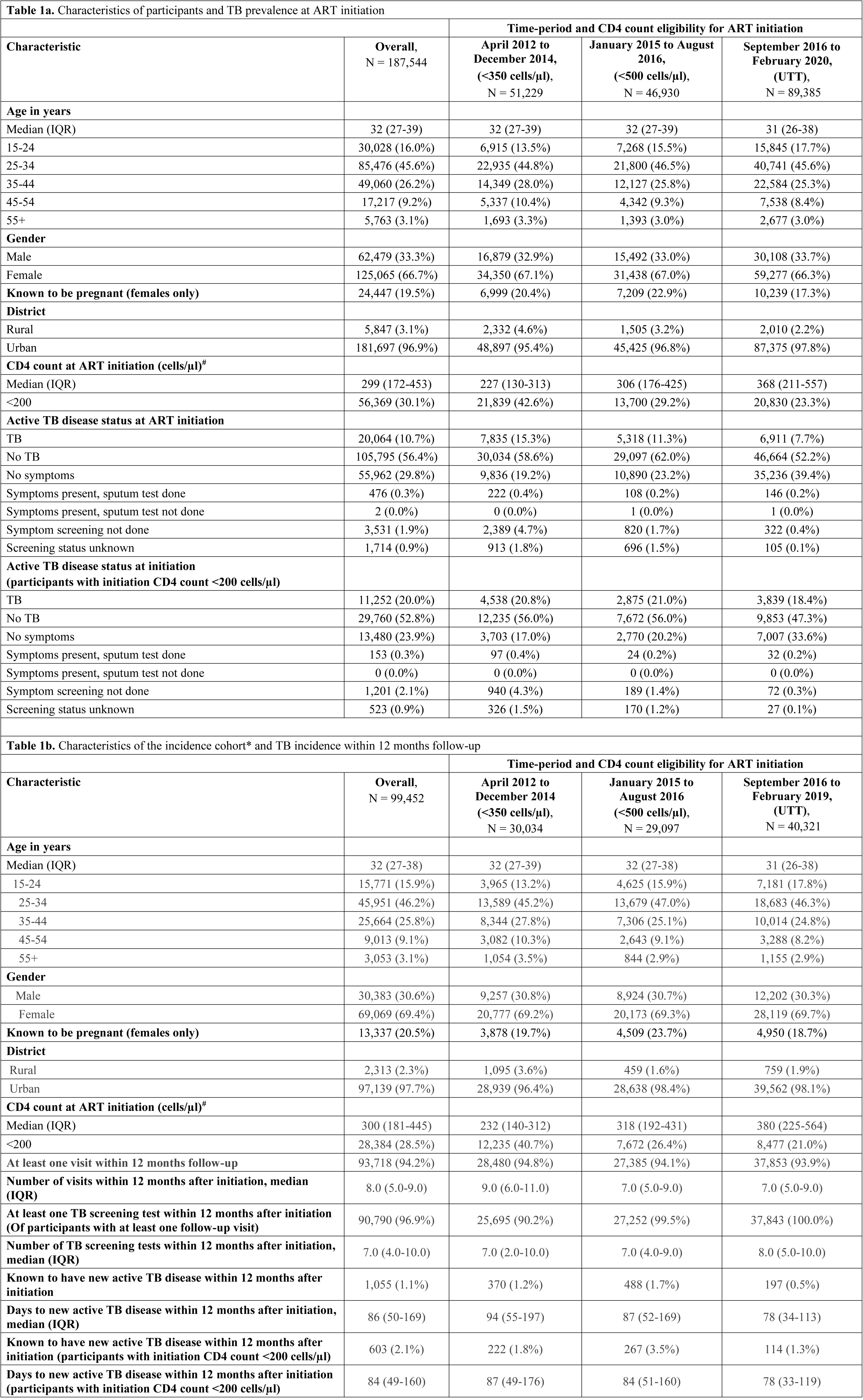
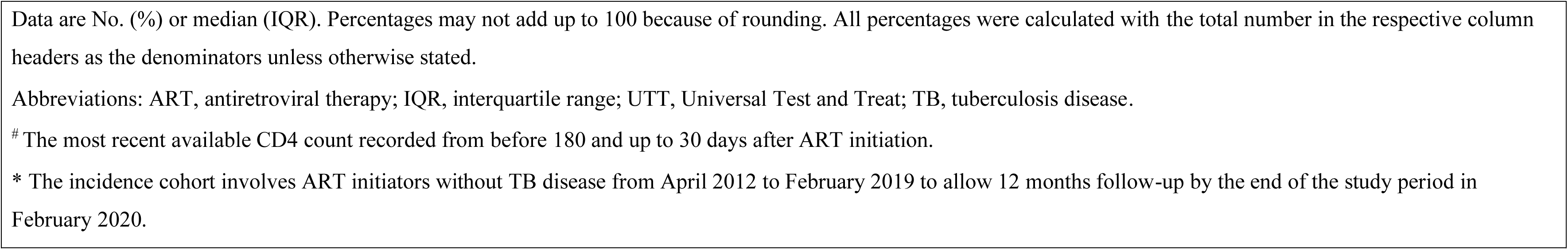

### Interrupted time series of new ART initiations from April 2012 to February 2020

From the segmented linear regression and interrupted time series analyses (Table 2A, Figure 1A), the monthly number of ART initiators was gradually rising, although with weak evidence of an increasing trend from April 2012 to December 2014 when the initiation CD4 count eligibility was <350 cells/μl. With the expansion of the initiation CD4 count eligibility to <500 cells/μl in January 2015, there was an immediate increase in the monthly number of ART initiators (Coeff 915.7, 95% confidence interval (CI) 422.3, 1409.1) and, thereafter, the slope decreased until August 2016 (Coeff -48.0, 95% CI -89.3, -6.7). From September 2016, when UTT was implemented, there was a further step increase in the monthly number of ART initiators (Coeff 832.9, 95% CI 369.9, 1295.9) with no evidence of a slope change until February 2020 (Coeff 3.7, 95% CI -34.8, 42.2). Among initiators with advanced HIV disease (Table 2A, Figure 1B), the monthly number of ART initiators was stable over time when the initiation CD4 count eligibility was <350 cells/μl and continued to remain stable with no step change (Coeff 77.2, 95% CI -33.1, 187.5) and no slope change (Coeff 0.0, 95% CI -8.6, 8.6) after the CD4 count eligibility was changed to <500 cells/μl. It further remained stable from September 2016, when UTT was implemented, with no evidence of a step change (Coeff 14.4, 95% CI -88.1 to 116.9) or slope change until February 2020 (Coeff -6.0, 95% CI -14.1, 2.1).

**Table 2.**
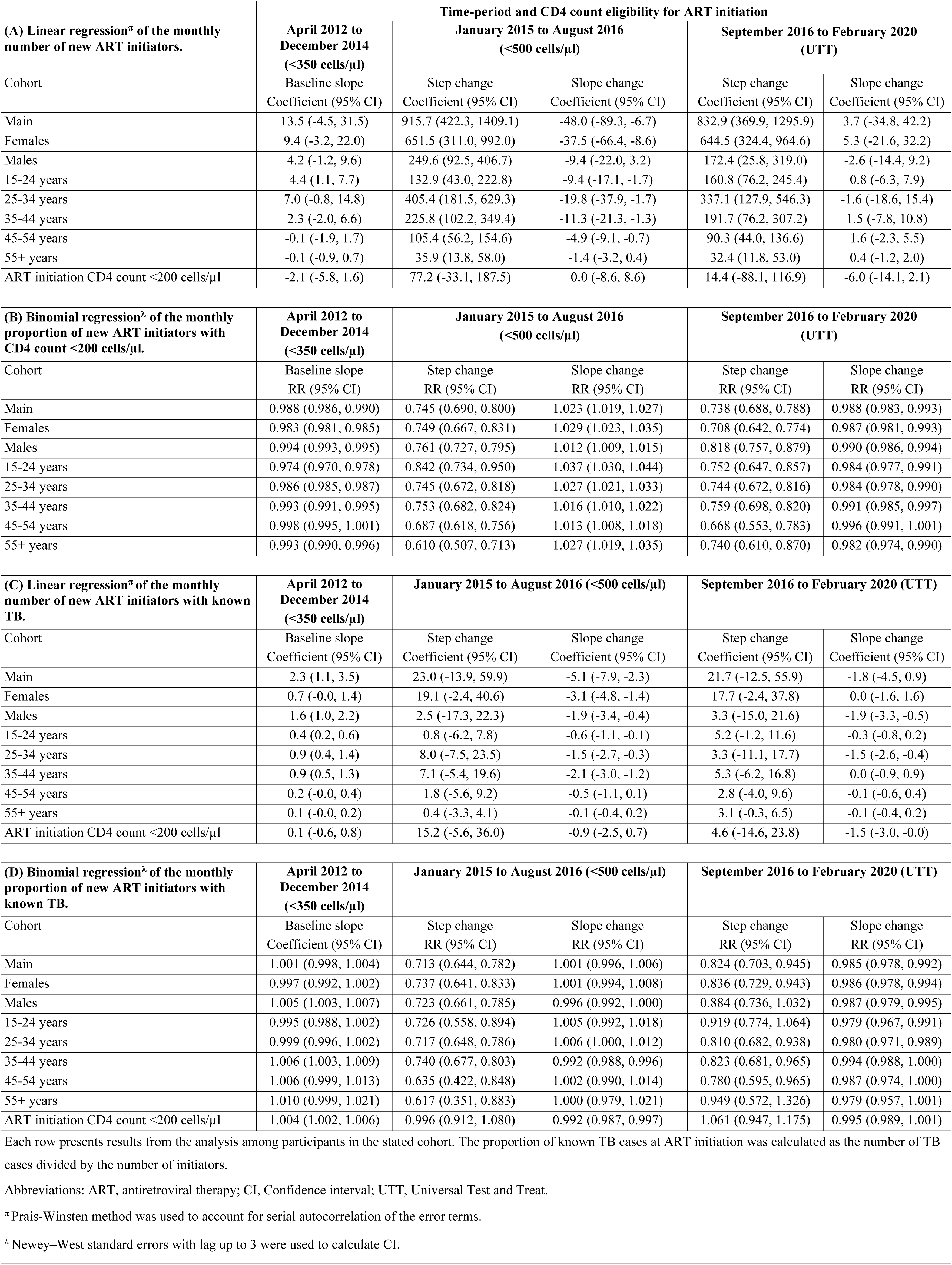
Impact of increasing CD4 count eligibility threshold for ART initiation on new ART initiations, advanced HIV disease at initiation and active TB disease prevalence at initiation from April 2012 to February 2020.

**Figure 1.**
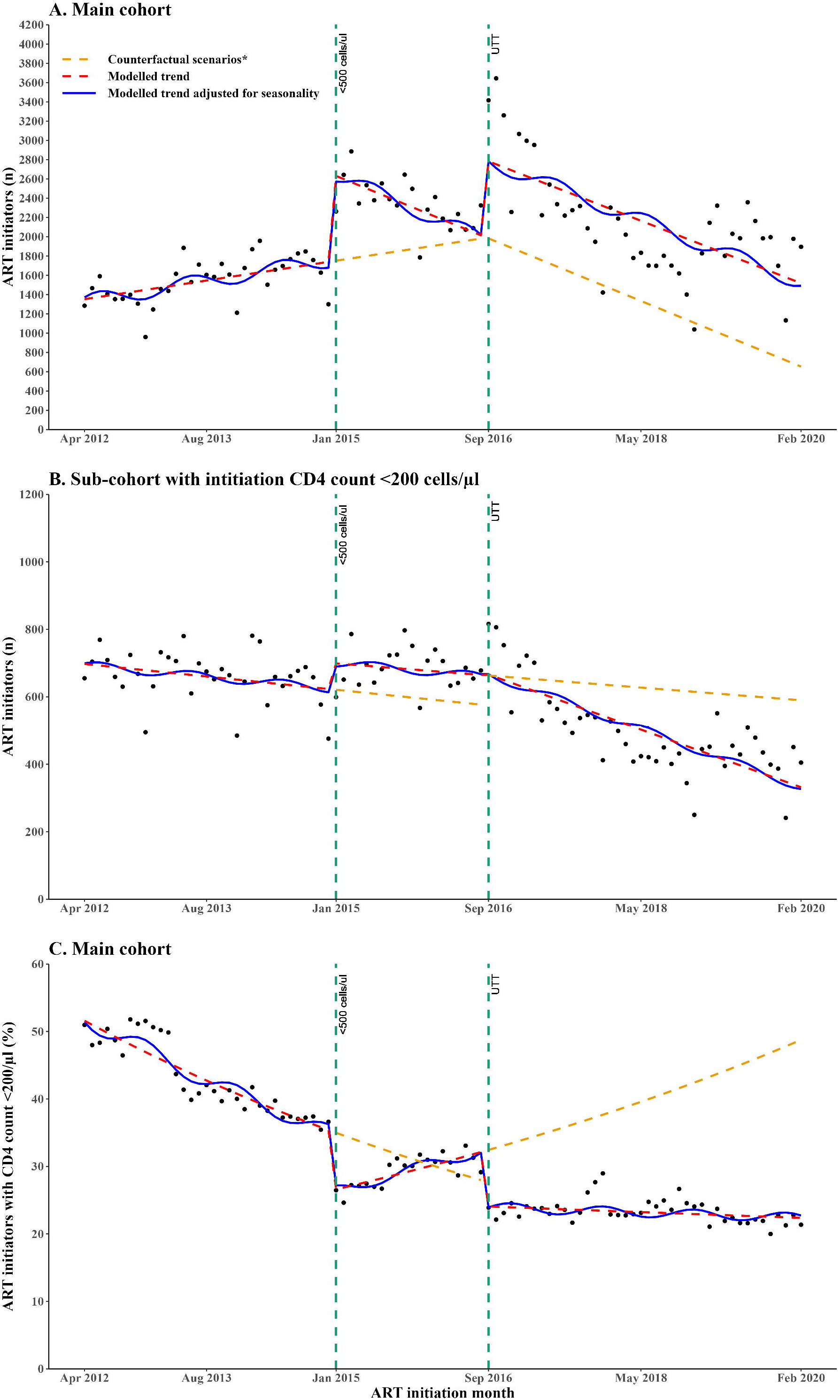
Segmented interrupted time series of (A) monthly number of new ART initiators, (B) monthly number of new ART initiators with advanced HIV disease and (C) monthly proportion of new ART initiators with advanced HIV disease. The time series spans three periods of different initiation CD4 count eligibility with two interruptions (vertical dashed lines) indicating the change points. Period 1 is from April 2012 to December 2014, when the initiation CD4 count eligibility was <350 cells/μl. Period 2 is from January 2015 to August 2016, when the initiation CD4 count eligibility was changed to <500 cells/μl. Period 3 is from September 2016 to February 2020 during UTT implementation. *Each counterfactual scenario represents the situation where only the previous initiation CD4 count expansion (s) was implemented. Abbreviations: ART, Antiretroviral therapy; PLHIV, people living with HIV; UTT, Universal Test and Treat.

### Interrupted time series of advanced HIV disease prevalence at ART initiation from April 2012 to February 2020

The segmented binomial regression and interrupted time series analyses of the proportion of initiators with advanced HIV disease are presented in Table 2B and Figure 1C. The results show a decreasing trend in the proportion of initiators with advanced HIV disease by 1.2% (Relative risk [RR] RR 0.988, 95% CI 0.986, 0.990) from April 2012 to December 2014 when the initiation CD4 count eligibility was <350 cells/μl. Afterwards, when the initiation CD4 count eligibility was expanded to <500 cells/μl in January 2015, there was a step decrease by 25.5% (RR 0.745, 95% CI 0.690, 0.800) and a slope decrease by 2.3% (RR 1.023, 95% CI 1.019, 1.027) until August 2016. Subsequently, when UTT was implemented in September 2016, there was a further step decrease by 26.2% (RR 0.738, 95% CI 0.688, 0.788) and a slope decrease by 1.2% until February 2020 (RR 0.988, 95% CI 0.983, 0.993). The differences in step and slope changes in the proportion of initiators with advanced HIV disease were similar by gender and by age group following expansion of the initiation CD4 count eligibility from <350 to <500 cells/μl and following UTT implementation.

### Interrupted time series of active TB disease prevalence at ART initiation from April 2012 to February 2020

From April 2012 to December 2014, when the initiation CD4 count eligibility was <350 cells/μl, the monthly number of initiators with TB was gradually rising (Table 2C, Figure 2A, Coeff 2.3, 95% CI 1.1, 3.5). There was no evidence of a step change in January 2015, when the initiation CD4 count eligibility was increased to <500 cells/μl (Coeff 23.0, 95% CI -13.9, 59.9). But afterwards, the slope decreased until August 2016 (Coeff -5.1, 95% CI -7.9, -2.3). There were no observed change in step but saw weak evidence of a slope decrease in the monthly number of initiators with TB after the introduction of UTT (Coeff -1.8, 95% CI -4.5, 0.9). Among initiators with advanced HIV disease (Table 2C, Figure 2B), the monthly number of initiators with TB was stable (Coeff 0.1, 95% CI -0.6, 0.8) when the initiation CD4 count eligibility was <350 cells/μl. Afterwards, there was no evidence of a step change (Coeff 15.2, 95% CI -5.6, 36.0) following the expansion of the initiation CD4 count eligibility to <500 cells/μl in January 2015 and no evidence of a slope change until August 2016 (Coeff - 0.9, 95% CI -2.5, 0.7). After UTT was implemented, there was no evidence of a step change (Coeff 4.6, 95% CI -14.6, 23.8) but weak evidence of a slope decrease until February 2020 (Coeff -1.5, 95% CI, -3.0, -0.0).

**Figure 2:**
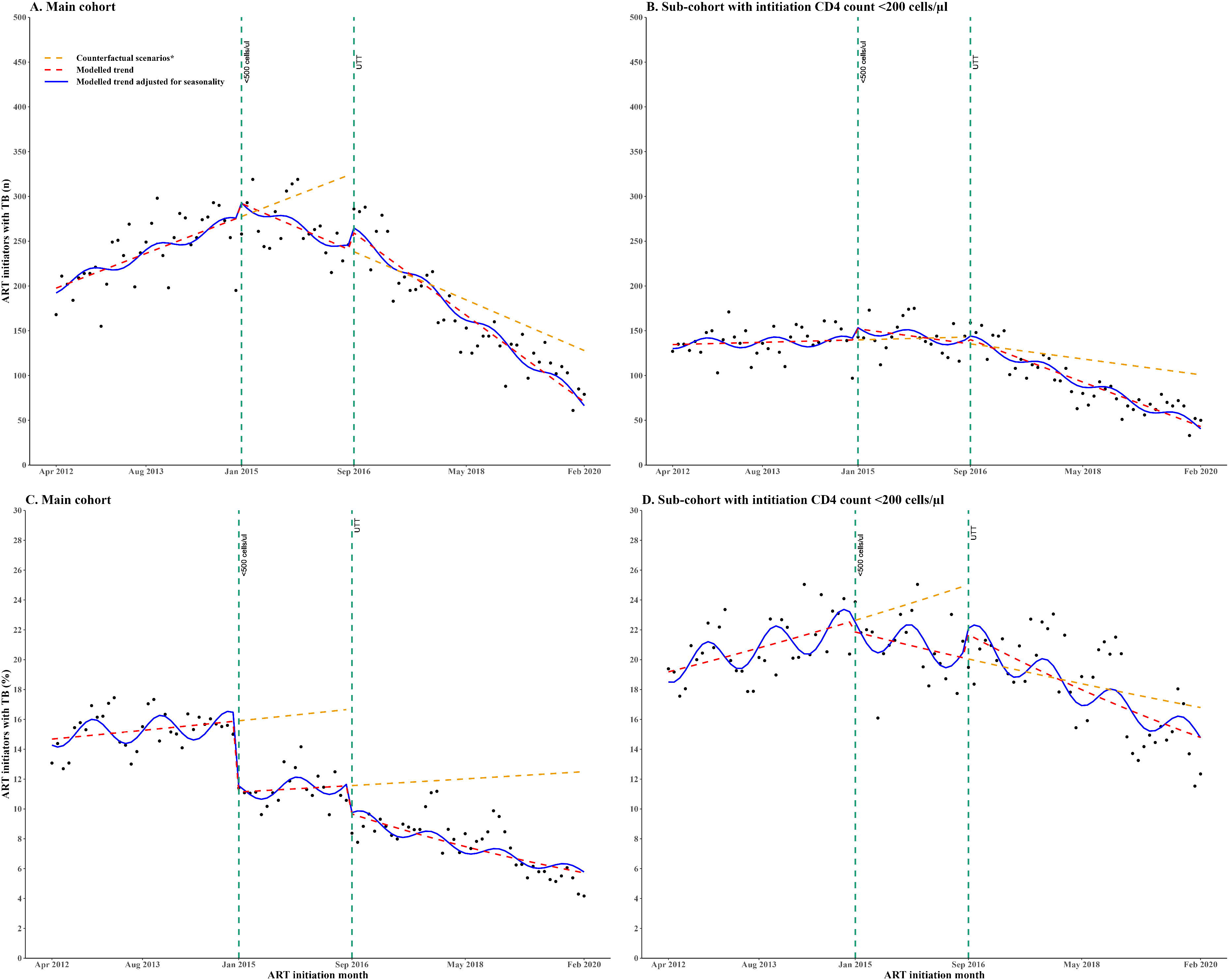
Segmented interrupted time series of (A) monthly number of new ART initiators with TB disease, (B) monthly number of new ART initiators with advanced HIV with TB disease, (C) monthly proportion of new ART initiators with TB disease, (D) monthly proportion of new ART initiators with advanced HIV with TB disease. The time series spans three periods of different initiation CD4 count eligibility with two interruptions (vertical dashed lines) indicating the change points. Period 1 is from April 2012 to December 2014, when the initiation CD4 count eligibility was <350 cells/μl. Period 2 is from January 2015 to August 2016, when the initiation CD4 count eligibility was changed to <500 cells/μl. Period 3 is from September 2016 to February 2020 during UTT implementation. *Each counterfactual scenario represents the situation where only the previous initiation CD4 count expansion was implemented. Abbreviations: ART, Antiretroviral therapy; TB, Tuberculosis; PLHIV, people living with HIV; UTT, Universal Test and Treat.

The monthly number of initiators with TB is a function of the number of people initiating ART (which changes over time), so we also present analyses of the proportion of initiators with TB. Table 2D and Figure 2C show that the proportion of initiators with TB was stable when the initiation CD4 count eligibility was <350 cells/μl. It then decreased by 28.7% (RR 0.713, 95% CI 0.644, 0.782) from January 2015 when the initiation CD4 count eligibility was expanded to <500 cells/μl but the slope remained stable until August 2016 (RR 1.001 95% CI 0.996, 1.006). Following UTT implementation in September 2016 there was a further step decrease of 17.6% (RR 0.824, 95% CI 0.703, 0.945) and a slope decline of 1.5% until February 2020 (RR 0.985 95% CI 0.978, 0.992). The proportion of initiators with TB across age groups and across gender were similar following the initiation CD4 count eligibility expansion from <350 to <500 cells/μl and following UTT implementation. Among initiators with advanced HIV disease (Table 2D and Figure 2D), the proportion with TB was stable over time when the initiation CD4 count eligibility was <350 cells/μl. With the expansion of the initiation CD4 count eligibility to <500 cells/μl, there was no evidence of a step change (RR 0.996, 95% CI 0.912, 1.080), but the slope decreased by 0.8% until August 2016 (RR 0.992, 95% CI 0.987, 0.997). Following the implementation of UTT from September 2016, there was no evidence of a step change (RR 1.061, 95% CI 0.947, 1.175) but weak evidence of a slope decrease until February 2020 (RR 0.995, 95% CI 0.989, 1.001).

### Characteristics of the incidence cohort and follow-up outcomes within 12 months

The incidence cohort involved 99,452 participants with no active TB disease at ART initiation from April 2012 to February 2019 (Table 1b). The median age was 32 years (IQR 27-38), and 69,069 (69.4%) were female, of whom 13,337 (20.5%) were pregnant at the time of ART initiation. The median CD4 count was 300 cells/μl (IQR 181-445), and 28,384 (28.5%) started ART with advanced HIV disease. The number and proportion with CD4 count <200 cells/μl was 12,235 (40.7%) when the initiation CD4 count eligibility was <350 cells/μl, 7,672 (26.4%) when the initiation CD4 count eligibility was <500 cells/μl and 8,477 (21.0%) during UTT implementation. The incidence cohort made a median of 8.0 (IQR, 5.0- 9.0) follow-up clinic visits within 12 months, and 93,718 (94.2%) made at least one follow-up clinic visit within the 12 months. Participants attending at least one follow-up clinic visit within 12 months received a median of 7.0 TB screening tests (IQR, 4.0-10.0) and 90,790 (96.9%) received at least one screening test for TB. The number and proportion of participants developing new TB within 12 months follow-up were 370 (1.2%) when the initiation CD4 count eligibility was <350 cells/μl, 488 (1.7%) when the initiation CD4 count eligibility was <500 cells/μl and 197 (0.5%) during UTT implementation. The median days to developing new TB within 12 months follow-up were 87 (49-176) when the initiation CD4 count eligibility was <350 cells/μl, 84 (51-160) when the initiation CD4 count eligibility was <500 cells/μl and 78 (33-119) during UTT implementation.

### Interrupted time series of the proportion of monthly ART initiators in the incidence cohort developing new TB within 12 months follow-up

Results from the segmented linear regression and interrupted time series analyses (Table 3A and Figure 3A) demonstrate no evidence of a changing trend in the number of initiators per month developing new TB within 12 months follow-up when the initiation CD4 count eligibility was <350 cells/μl. Following the expansion of the initiation CD4 count eligibility to <500 cells/μl in January 2015, there was a step increase (Coeff 11.9, 95% CI 7.0, 16.8) but no evidence of a slope change until August 2016 (Coeff -0.2, 95% CI -0.6, 0.2). With the implementation of UTT from September 2016, there was a step decrease (Coeff -6.9, 95% CI -11.7, -2.1) and a slope decrease until February 2019 (Coeff -0.6, 95% -1.0, -0.2). Among the incidence cohort with advanced HIV disease (Table 3A and Figure 3B), the number developing new TB within 12 months of follow-up exhibited a stable trend when the initiation CD4 count eligibility was <350 cells/μl. Following the expansion of the initiation CD4 count eligibility to <500 cells/μl in January 2015, there was weak evidence of a step increase (Coeff 2.1, 95% CI -1.1, 5.3) and weak evidence of a slope increase until August 2016 (Coeff 0.3, 95% CI 0.1, 0.5). However, following UTT implementation from September 2016, there was a step decrease (Coeff -7.7, 95% CI -11.1, -4.3) and a slope decrease until February 2019 (Coeff -0.7, 95% CI, -0.7, -0.9).

**Table 3.**
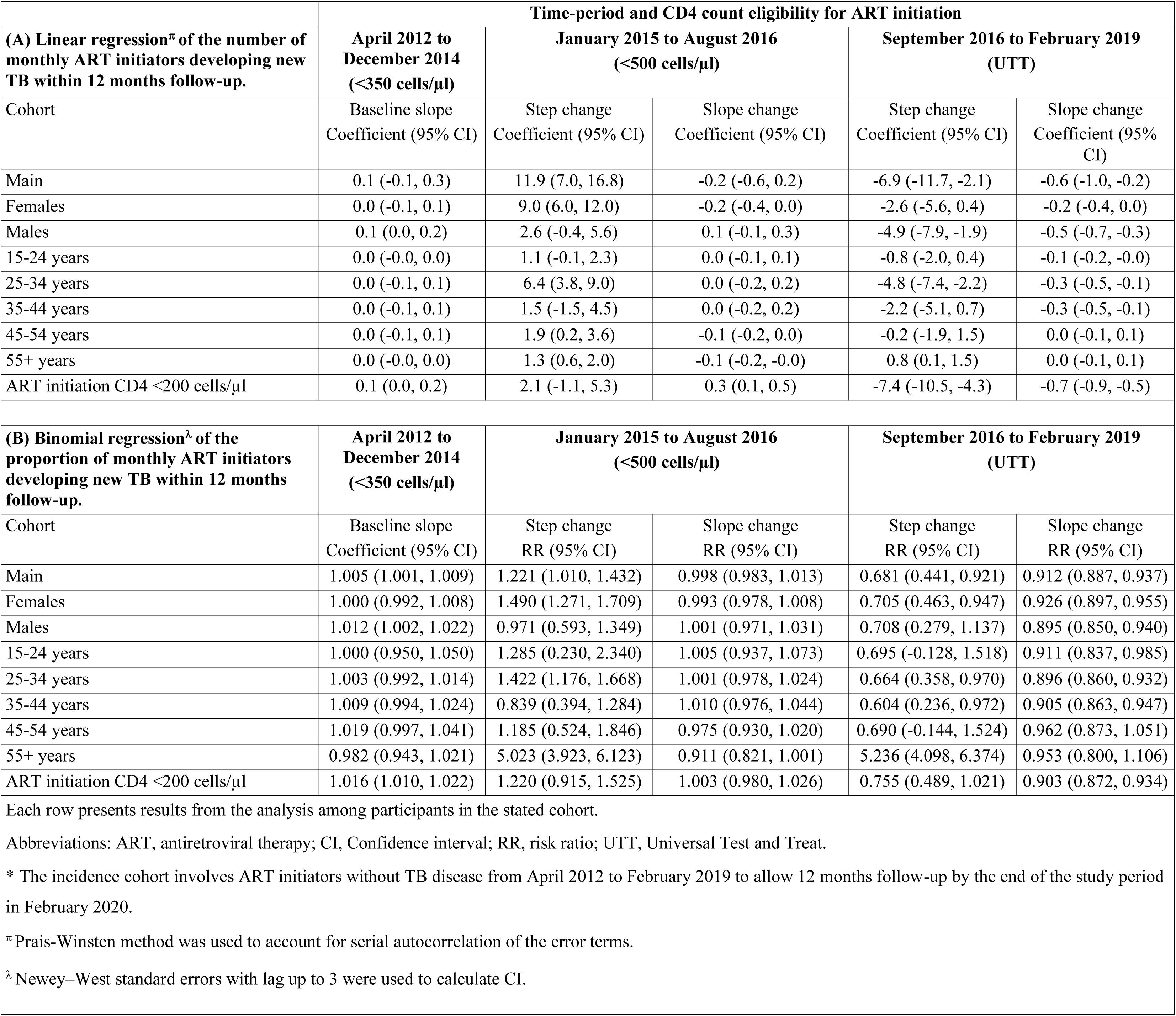
Impact of increasing CD4 count eligibility threshold for ART initiation on cumulative TB incidence within 12 months follow-up among the incidence cohort.

**Figure 3.**
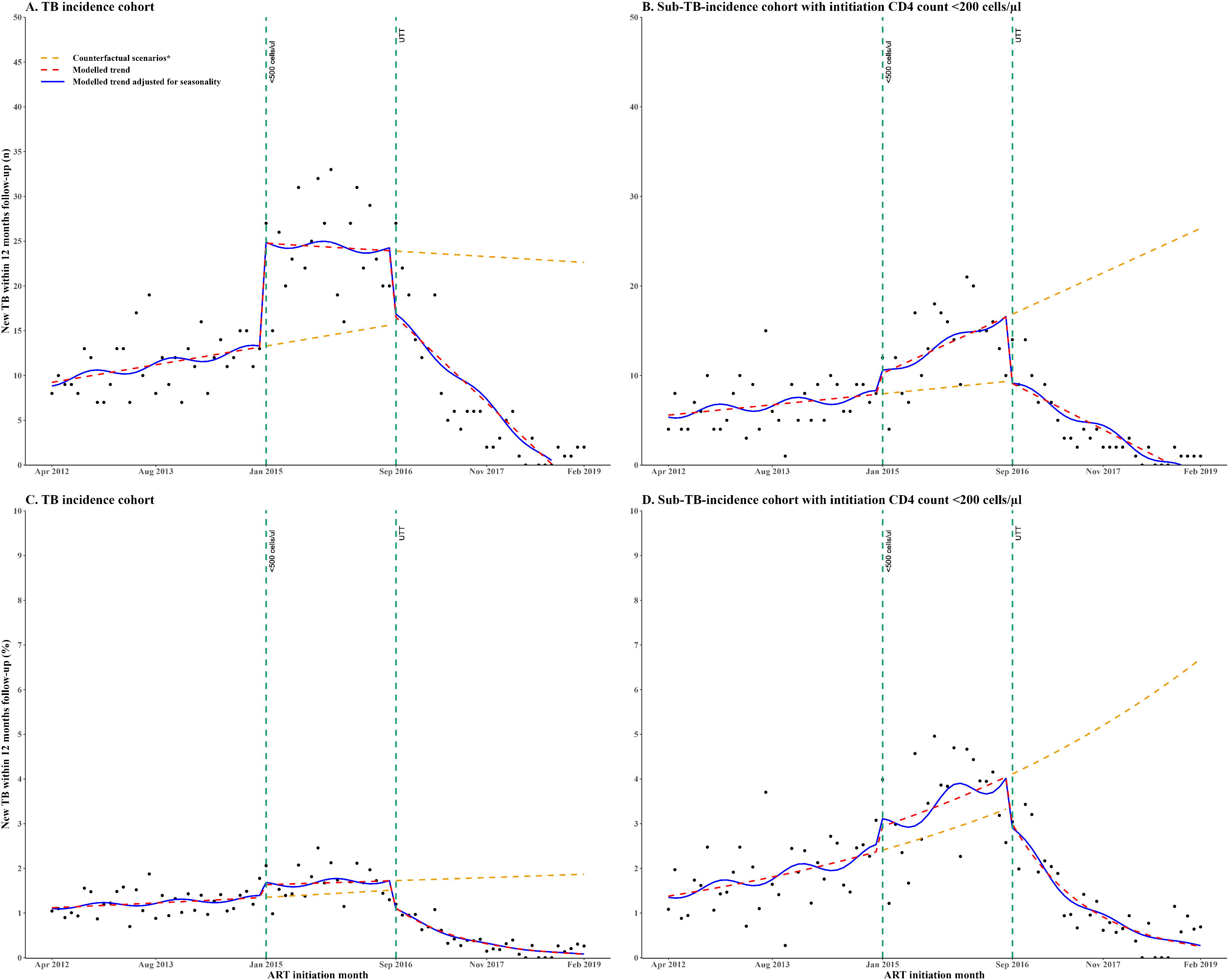
Segmented interrupted time series among the incidence cohort of initiators without TB disease: (A) monthly number developing new TB disease within 12 months follow-up, (B) monthly number with advanced HIV developing new TB disease within 12 months follow-up, (C) monthly proportion developing new TB disease within 12 months follow-up and (D) monthly proportion with advanced HIV developing new TB disease within 12 months follow-up. The time series spans three periods of different initiation CD4 count eligibility with two interruptions (vertical dashed lines) indicating the change points. Period 1 is from April 2012 to December 2014, when the initiation CD4 count eligibility was <350 cells/μl. Period 2 is from January 2015 to August 2016, when the initiation CD4 count eligibility was changed to <500 cells/μl. Period 3 is from September 2016 to February 2019 during UTT implementation. *Each counterfactual scenario represents the situation where only the previous initiation CD4 count expansion was implemented. Abbreviations: ART, Antiretroviral therapy; TB, Tuberculosis; PLHIV, people living with HIV; UTT, Universal Test and Treat

Results from the segmented binomial regression and interrupted time series analyses (Table 3B and Figure 3C) demonstrate that the proportion of monthly initiators developing new TB within 12 months of follow-up was stable when the initiation CD4 count was <350 cells/μl. When the initiation CD4 count was increased to <500 cells/μl in January 2015, there was no evidence of a step change (RR 1.221, 95% CI 1.010, 1.432) and no evidence of a slope change until August 2016 (RR 0.998, 95% CI 0.983, 1.013). However, following the implementation of UTT in September 2016, there was a step decrease by 31.9% (RR 0.681, 95% CI 0.441, 0.921) and a slope decrease by 8.8% until February 2019 (RR 0.912, 95% CI 0.887, 0.937). Among the incidence cohort with advanced HIV disease, the results (Table 3B and Figure 3D) show no evidence of a changing trend in the proportion of monthly initiators developing new TB within 12 months follow-up when the initiation CD4 count was <350 cells/μl. Subsequently, when the initiation CD4 count eligibility was expanded to <500 in January 2015, there was no evidence of a step change (RR 1.220, 95% CI 0.915, 1.525) and no evidence of a slope change until August 2016 (RR 1.003, 95% CI 0.980 to 1.026).

However, from September 2016, when UTT was implemented, there was weak evidence of a step change (RR 0.755 95% CI 0.489, 1.021), and the slope decreased by 9.7% until February 2019 (RR 0.903, 95% CI 0.872, 0.934).

## DISCUSSION

We present results from an interrupted time series analysis of de-identified large-scale routinely collected clinic data from South Africa’s ART programme between April 2012 and February 2020. The findings show that, following the expansion of the ART initiation CD4 count eligibility from <350 to <500 cells/μl in January 2015 and the subsequent implementation of UTT in September 2016, ART initiations immediately increased but thereafter declined approaching the counterfactual levels associated with previous CD4 count eligibility criteria. The declining trend in new ART initiations was partly due to the existing backlog of ART naïve PLHIV who immediately became eligible for initiation, that gradually cleared over time. Additionally, the monthly proportion of ART initiators with advanced HIV disease decreased although about a quarter of people still started ART with advanced HIV disease after increasing the CD4 eligibility for ART initiation to <500 cells/μl in January 2015 and during the UTT era. The pooled prevalence of advance HIV disease among ART-naïve patients from a systematic review of 53 studies conducted between 2010 to 2022 (majority before 2016) was 43.4% (95% CI 40.1–46.8%). This pooled prevalence is consistent with the prevalence of advanced HIV disease in our study from April 2012 to January 2015 during when the CD4 count eligibility for ART initiation was <350 cells/μl.

Additionally, our estimate of advance HIV disease in the UTT era is consistent with results from the studies that were conducted from 2016 onwards.[29, 30] We also found that TB prevalence among all initiators declined following expansion of CD4 count eligibility criteria but remained stable among the subset of initiators with advanced HIV disease. Finally, among initiators without TB disease, the cumulative incidence of TB within 12 months of follow-up declined following UTT implementation, in the whole cohort and in the cohort of individuals that initiated with advanced HIV disease. Generally, the impact of expanded ART access on the prevalence of advanced HIV disease at initiation, TB prevalence at initiation and TB incidence within 12 months follow-up were similar by gender and age.

Some studies have previously assessed the impact of expanded initiation CD4 count eligibility on new ART initiations, advanced HIV disease and TB disease burden among people initiating and receiving ART from routine care settings.[9, 14–16, 18] Results from a cohort study among adult participants in the rural Hlabisa HIV treatment programme in KwaZulu-Natal, South Africa, demonstrated increased new ART initiations in PLHIV but did not change among initiators with advanced HIV disease following expansion of the initiation CD4 count eligibility from <200 to <350 cells/μl in August 2011.[15] Other studies in South Africa have reported higher mean initiation CD4 count[8] and a lower proportion of initiators with advanced HIV disease[9] after UTT implementation than before. Additionally, the African cohort study in four countries reported increased new ART initiations at higher initiation CD4 count following the implementation of policies and guidelines that expanded initiation CD4 count eligibility in the selected countries from 2006 to 2019.[31]

Regarding the impact of expanded ART access on TB burden, our results are consistent with a large scale study in South Africa that showed lower risk of recently diagnosed TB cases in communities with higher ART coverage.[18] We found a cluster randomised trial in Uganda that evaluated a UTT intervention involving population-level HIV testing and patient-centred linkage to ART versus population-level HIV testing in the control communities. At the end of the trial, the one-year cumulative incidence of TB infection was 16% in the intervention and 22% in the control communities.[14] Likewise, the risk of new TB infection was 27% lower with the UTT intervention.[14] Another study investigated the impact of the UTT policy on TB incidence in a cohort of adults receiving ART in public health facilities in Ethiopia from 2014 to 2019.[16] The study reported TB incidence of 6.23 cases per 100 person-years before UTT implementation versus 2.10 cases per 100 person-years after UTT.[16] Furthermore, estimates from WHO’s 2022 global tuberculosis report have shown a declining trend in the population-level incidence of TB co-burden in PLHIV in the WHO African Region[32] and in South Africa[33] after 2010.

We used large-scale data from routine ART clinics in a resource-limited setting. Therefore, our results are representative and relevant for ART programmes in resource-limited settings. The findings are reassuring of the positive impact of expanded ART access for reducing the risk of advanced HIV disease and opportunistic co-infections which could improve the health and well-being of PLHIV. Our findings indicate that extending the initiation CD4 count from <350 to <500 cells/μl in January 2015 allowed more PLHIV to start ART than previously. Later on, removing the initiation CD4 count testing requirement and eligibility criteria with UTT implementation saw further decreases in new ART initiations. However, our results show that despite these gains, about 23.3% of PLHIV still presented with advanced HIV disease at initiation, although this likely underestimates advanced HIV disease prevalence as our definition was only based on CD4 count threshold. People who initiate ART with advanced HIV disease in the UTT era are more likely to have missed the opportunity for early HIV diagnosis. Our findings, therefore, emphasise the continuing urgency for health systems in resource-limited settings to prioritise access to routine HIV testing services, especially among higher-risk populations such as younger people, pregnant women and their male partners. This will help facilitate early HIV diagnosis and immediate linkage to care to exploit the full potential of UTT policy implementation.

Moreover, our study demonstrates the positive impact of expanded ART access on TB co-burden in PLHIV. Our findings suggest that people initiating ART currently in the UTT era are less likely to present with TB. This is due to more people initiating ART at higher CD4 counts with more robust immune functions[15, 34, 35], reducing the risk of opportunistic co-infections such as TB.[36–38] Our findings also indicate that people initiating ART without TB in the UTT era are less likely to develop new TB while on ART compared to the pre-UTT period. This could be attributed to early ART initiation[15, 34, 35], ensuring early viral suppression, which enhances consistent and prolonged immune functioning.[11, 39, 40] Furthermore, we found a trend towards a decrease in TB incidence among initiators with advanced HIV disease. This suggests the positive impact of expanded ART access on TB transmission risk among PLHIV due to reduced exposure to TB bacterium as fewer people initiate ART without TB. We did not observe any impact of expanding the initiation CD4 count eligibility from <350 to <500 cells/μl in January 2015 on decreased TB disease incidence but this could be due to the shorter duration of the criteria change (21 months) before UTT implementation in September 2016. It is therefore likely that the decreased TB incidence observed after UTT implementation represents the cumulative impact of the overall expanded ART access over time (all initiation CD4 count criteria expansions).

Our study had some limitations. Although our data set was large, it came from only one province in South Africa. We, therefore, acknowledge that outcomes could be different from other settings. Also, data quality, correctness and completion rates in TIER.Net has improved overtime compared to the early years after implementation in 2010 which could bias our results.[41] We also acknowledge the potential confounding of our study outcomes by other concurrent interventions that could have contributed to better outcomes in the UTT era such as more efficacious and tolerable ART regimens and improvements in TB preventive therapy.[42, 43] Therefore, participants who initiated ART recently or during the UTT era are more likely to have initiated more effective and tolerable regimens. Hence, they are more likely to be retained in care[44], virally suppress[45, 46] and consequently less likely to develop new TB during ART. Moreover, the implementation of Central Chronic Medication Dispensing and Distribution programme in Kwazulu-Natal in 2016[47] could have reduced patient volumes and hence reduced exposure to TB at the clinics contributing to the declining TB burden we observed in the UTT era.

In conclusion, expansion of the CD4 count eligibility for ART initiation over time has led to expanded access to ART and a healthier population of PLHIV with stronger immunity and reduced risk for co-TB disease. Health systems in resource-limited and HIV-endemic settings should prioritise population-level early HIV diagnosis to maximise the full potential of UTT policy implementation. Tailored interventions focusing on people with advance HIV disease at ART initiation and during ART are also needed to improve treatment outcomes and reduce opportunistic co-infections like TB disease.

## Acknowledgements

We acknowledge staff, patients and primary care clinics run by the eThekwini Municipality Health Unit and Bethesda Hospital in the uMkhanyakude district in South Africa.

## Contributors

The study was conceptualised by KA, LL, JvdM, YS, TK, TN, MSN, RJL, KN, PS, CB, NG, and JD. YS, TK, TN, MSN and PS managed data collection. KA, LL, JvdM, YS, TK, TN, MSN, NG and JD had full access to the data in the study. KA, LL, JvdM, and JD conducted the data analyses. KA drafted the manuscript. All authors reviewed and approved the final version.

## Funding

This study was supported with funding from the Bill & Melinda Gates Foundation (INV-051067). JD is an academic clinical lecturer (CL-2022-13-005) funded by the UK National Institute of Health and Social Care Research (NIHR). The views expressed in this publication are those of the author (s) and not necessarily those of the Bill & Melinda Gates Foundation or NIHR.

## Competing interest

None declared.

## Patient consent for publication

Not required.

## Ethics approval

The study was approved by the Biomedical Research Ethics Committee of the University of Kwazulu-Natal (BE646/17), the KwaZulu-Natal Provincial Health Research Ethics Committee (KZ_201807_021), the eThekwini Municipality Health Unit and the Bethesda Hospital Ethics Committee, with a waiver for informed consent for the analysis of de-identified data.

## Data availability statement

We cannot publicly share the data used for this study due to legal and ethical requirements regarding using routinely collected clinical data in South Africa. Interested parties can request access to the data from the eThekwini Municipality Health Unit in Durban and Bethesda Hospital in the uMkhanyakude District, South Africa.

## Notes

### Competing Interest Statement

The authors have declared no competing interest.

## REFERENCES

1. Govere SM, Chimbari MJ. The evolution and adoption of World Health Organization policy guidelines on antiretroviral therapy initiation in sub-Saharan Africa: A scoping review. Southern African Journal of HIV Medicine. 2020;21(1).

2. South African National Department of Health. Implementation of universal test and treat strategy for HIV positive patients and differentiated care for stable patients. 2016 [Available from: https://sahivsoc.org/Files/22%208%2016%20Circular%20UTT%20%20%20Decongestion%20CCMT%20Directorate.pdf.

3. World Health Organisation. Progress Report 2016. Prevent HIV, Test and Treat All. WHO support for country impact. 2016 [Available from: https://apps.who.int/iris/bitstream/handle/10665/251713/WHO-HIV-2016.24-eng.pdf.

4. South African Government. National Strategic Plan on HIV, STIs and TB 2012-2016. 2012 [Available from: https://www.gov.za/sites/default/files/gcis_document/201409/national-strategic-plan-hiv-stis-and-tb0.pdf.

5. Cohen MS, Chen YQ, Mccauley M, Gamble T, Hosseinipour MC, Kumarasamy N, et al. Prevention of HIV-1 Infection with Early Antiretroviral Therapy. New England Journal of Medicine. 2011;365(6):493–505.

6. Abdool Karim SS, Naidoo K, Grobler A, Padayatchi N, Baxter C, Gray A, et al. Timing of initiation of antiretroviral drugs during tuberculosis therapy. N Engl J Med. 2010;362(8):697–706.

7. Kaplan R, Hermans S, Caldwell J, Jennings K, Bekker L-G, Wood R. HIV and TB co-infection in the ART era: CD4 count distributions and TB case fatality in Cape Town. BMC Infectious Diseases. 2018;18(1).

8. Yapa HM, Kim HY, Petoumenos K, Post FA, Jiamsakul A, De Neve JW, et al. CD4+ T-Cell Count at Antiretroviral Therapy Initiation in the “Treat-All” Era in Rural South Africa: An Interrupted Time Series Analysis. Clin Infect Dis. 2022;74(8):1350–9.

9. Zaniewski E, Dao Ostinelli CH, Chammartin F, Maxwell N, Davies MA, Euvrard J, et al. Trends in CD4 and viral load testing 2005 to 2018: multi-cohort study of people living with HIV in Southern Africa. J Int AIDS Soc. 2020;23(7):e25546.

10. Filiatreau LM, Edwards JK, Masilela N, Gómez-Olivé FX, Haberland N, Pence BW, et al. Understanding the effects of universal test and treat on longitudinal HIV care outcomes among South African youth: a retrospective cohort study. BMC Public Health. 2023;23(1).

11. Hoenigl M, Chaillon A, Moore DJ, Morris SR, Mehta SR, Gianella S, et al. Rapid HIV Viral Load Suppression in those Initiating Antiretroviral Therapy at First Visit after HIV Diagnosis. Scientific Reports. 2016;6(1):32947.

12. Johnson LF, Meyer-Rath G, Dorrington RE, Puren A, Seathlodi T, Zuma K, et al. The Effect of HIV Programs in South Africa on National HIV Incidence Trends, 2000–2019. JAIDS Journal of Acquired Immune Deficiency Syndromes. 2022;90(2):115–23.

13. Burger C, Burger R, Van Doorslaer E. The health impact of free access to antiretroviral therapy in South Africa. Social Science & Medicine. 2022;299:114832.

14. Marquez C, Atukunda M, Nugent J, Charlebois ED, Chamie G, Mwangwa F, et al. Community-Wide Universal Human Immunodeficiency Virus (HIV) Test and Treat Intervention Reduces Tuberculosis Transmission in Rural Uganda: A Cluster-Randomized Trial. Clinical Infectious Diseases. 2024.

15. Plazy M, Dabis F, Naidu K, Orne-Gliemann J, Barnighausen T, Dray-Spira R. Change of treatment guidelines and evolution of ART initiation in rural South Africa: data of a large HIV care and treatment programme. BMC Infectious Diseases. 2015;15(1).

16. Girum T, Yasin F, Dessu S, Zeleke B, Geremew M. “Universal test and treat” program reduced TB incidence by 75% among a cohort of adults taking antiretroviral therapy (ART) in Gurage zone, South Ethiopia. Tropical Diseases, Travel Medicine and Vaccines. 2020;6(1).

17. Kitenge MK, Fatti G, Eshun-Wilson I, Aluko O, Nyasulu P. Prevalence and trends of advanced HIV disease among antiretroviral therapy-naive and antiretroviral therapy-experienced patients in South Africa between 2010-2021: a systematic review and meta-analysis. BMC Infect Dis. 2023;23(1):549.

18. Tomita A, Smith CM, Lessells RJ, Pym A, Grant AD, de Oliveira T, et al. Space-time clustering of recently-diagnosed tuberculosis and impact of ART scale-up: Evidence from an HIV hyper-endemic rural South African population. Sci Rep. 2019;9(1):10724.

19. The South African National Department of Health. 2019 ART Clinical Guidelines for the management of HIV in Adults, Pregnancy, Adolescents, Children, Infants and Neonates. Pretoria2019 [Available from: https://www.health.gov.za/wp-content/uploads/2020/11/2019-art-guideline.pdf.

20. The South African National Department of Health. The South African Antiretroviral Treatment Guidelines. 2013 [Available from: https://sahivsoc.org/Files/2013%20ART%20Treatment%20Guidelines%20Final%2025%20March%202013%20corrected.pdf.

21. Churchyard GJ, Stevens WS, Mametja LD, McCarthy KM, Chihota V, Nicol MP, et al. Xpert MTB/RIF versus sputum microscopy as the initial diagnostic test for tuberculosis: a cluster-randomised trial embedded in South African roll-out of Xpert MTB/RIF. Lancet Glob Health. 2015;3(8):e450–e7.

22. Dorward J, Khubone T, Gate K, Ngobese H, Sookrajh Y, Mkhize S, et al. The impact of the COVID-19 lockdown on HIV care in 65 South African primary care clinics: an interrupted time series analysis. Lancet HIV. 2021;8(3):e158–e65.

23. Osler M, Hilderbrand K, Hennessey C, Arendse J, Goemaere E, Ford N, et al. A three-tier framework for monitoring antiretroviral therapy in high HIV burden settings. Journal of the International AIDS Society. 2014;17(1):18908.

24. Bottomley C, Ooko M, Gasparrini A, Keogh RH. In praise of Prais-Winsten: An evaluation of methods used to account for autocorrelation in interrupted time series. Stat Med. 2023;42(8):1277–88.

25. Bernal JL, Cummins S, Gasparrini A. Interrupted time series regression for the evaluation of public health interventions: a tutorial. Int J Epidemiol. 2017;46(1):348–55.

26. Bottomley C, Scott JAG, Isham V. Analysing Interrupted Time Series with a Control. Epidemiologic Methods. 2019;8(1).

27. Wagner AK, Soumerai SB, Zhang F, Ross-Degnan D. Segmented regression analysis of interrupted time series studies in medication use research. Journal of Clinical Pharmacy and Therapeutics. 2002;27(4):299–309.

28. R Core Team. R: A language and environment for statistical computing Vienna, Austria: R Foundation for Statistical Computing; 2023 [Available from: https://www.R-project.org/.

29. Cassidy T, Cornell M, Makeleni B, Horsburgh CR, Duran LT, de Azevedo V, et al. Attrition from Care Among Men Initiating ART in Male-Only Clinics Compared with Men in General Primary Healthcare Clinics in Khayelitsha, South Africa: A Matched Propensity Score Analysis. AIDS Behav. 2023;27(1):358–69.

30. Bock P, Fatti G, Ford N, Jennings K, Kruger J, Gunst C, et al. Attrition when providing antiretroviral treatment at CD4 counts >500cells/muL at three government clinics included in the HPTN 071 (PopART) trial in South Africa. PLoS One. 2018;13(4):e0195127.

31. Esber AL, Coakley P, Ake JA, Bahemana E, Adamu Y, Kiweewa F, et al. Decreasing time to antiretroviral therapy initiation after HIV diagnosis in a clinic-based observational cohort study in four African countries. J Int AIDS Soc. 2020;23(2):e25446.

32. World Health Organisation. Global tuberculosis report 2022. 2022 [Available from: https://www.who.int/publications/i/item/9789240061729.

33. World Health Organisation. Tuberculosis profile: South Africa. 2022 [Available from: https://worldhealthorg.shinyapps.io/tb_profiles/?_inputs_&entity_type=%22country%22&iso2=%22ZA%22&lan=%22EN%22.

34. Bor J, Ahmed S, Fox MP, Rosen S, Meyer-Rath G, Katz IT, et al. Effect of eliminating CD4-count thresholds on HIV treatment initiation in South Africa: An empirical modeling study. PLOS ONE. 2017;12(6):e0178249.

35. Russell A, Verani AR, Pals S, Reagon VM, Alexander LN, Galloway ET, et al. Impact of HIV treat-all and complementary policies on ART linkage in 13 PEPFAR-supported African countries. BMC Health Services Research. 2023;23(1).

36. Lawn SD, Kranzer K, Wood R. Antiretroviral therapy for control of the HIV-associated tuberculosis epidemic in resource-limited settings. Clin Chest Med. 2009;30(4):685–99, viii.

37. McLaren ZM, Sharp A, Brouwer E, Nanoo A. The Impact of Anti-Retroviral Therapy on Tuberculosis Detection at the National Level in South Africa. Am J Trop Med Hyg. 2018;99(6):1407–14.

38. Suthar AB, Lawn SD, Del Amo J, Getahun H, Dye C, Sculier D, et al. Antiretroviral Therapy for Prevention of Tuberculosis in Adults with HIV: A Systematic Review and Meta-Analysis. PLoS Medicine. 2012;9(7):e1001270.

39. Dorward J, Sookrajh Y, Gate K, Khubone T, Mtshaka N, Mlisana K, et al. HIV treatment outcomes among people with initiation CD4 counts >500 cells/µL after implementation of Treat All in South African public clinics: a retrospective cohort study. J Int AIDS Soc. 2020;23(4):e25479.

40. Eamsakulrat P, Kiertiburanakul S. The Impact of Timing of Antiretroviral Therapy Initiation on Retention in Care, Viral Load Suppression and Mortality in People Living with HIV: A Study in a University Hospital in Thailand. J Int Assoc Provid AIDS Care. 2022;21:23259582221082607.

41. Etoori D, Wringe A, Kabudula CW, Renju J, Rice B, Gomez-Olive FX, et al. Misreporting of Patient Outcomes in the South African National HIV Treatment Database: Consequences for Programme Planning, Monitoring, and Evaluation. Front Public Health. 2020;8:100.

42. Kanters S, Vitoria M, Zoratti M, Doherty M, Penazzato M, Rangaraj A, et al. Comparative efficacy, tolerability and safety of dolutegravir and efavirenz 400mg among antiretroviral therapies for first-line HIV treatment: A systematic literature review and network meta-analysis. EClinicalMedicine. 2020;28:100573.

43. World Health Organisation. WHO consolidated guidelines on tuberculosis: tuberculosis preventive treatment: Module 1: prevention Geneva: World Health Organization; 2020 [

44. Alhaj M, Amberbir A, Singogo E, Banda V, van Lettow M, Matengeni A, et al. Retention on antiretroviral therapy during Universal Test and Treat implementation in Zomba district, Malawi: a retrospective cohort study. J Int AIDS Soc. 2019;22(2):e25239.

45. Dorward J, Sookrajh Y, Khubone T, Van Der Molen J, Govender R, Phakathi S, et al. Implementation and outcomes of dolutegravir-based first-line antiretroviral therapy for people with HIV in South Africa: a retrospective cohort study. The Lancet HIV. 2023.

46. Asare K, Sookrajh Y, van der Molen J, Khubone T, Lewis L, Lessells RJ, et al. Clinical outcomes with second-line dolutegravir in people with virological failure on first-line non-nucleoside reverse transcriptase inhibitor-based regimens in South Africa: a retrospective cohort study. Lancet Glob Health. 2023.

47. KwaZulu-Natal Department of Health. Launch of the Central Chronic Medication Dispensing and Distribution programme. 2016 [Available from: https://www.kznhealth.gov.za/Launch-CCMDD-10062016.htm.

